# Association of social distancing and masking with risk of COVID-19

**DOI:** 10.1101/2020.11.11.20229500

**Authors:** Sohee Kwon, Amit D. Joshi, Chun-Han Lo, David A. Drew, Long H. Nguyen, Chuan-Guo Guo, Wenjie Ma, Raaj S. Mehta, Erica T. Warner, Christina M. Astley, Jordi Merino, Benjamin Murray, Jonathan Wolf, Sebastien Ourselin, Claire J. Steves, Tim D. Spector, Jaime E. Hart, Mingyang Song, Trang VoPham, Andrew T. Chan

## Abstract

Given the continued burden of severe acute respiratory syndrome coronavirus 2 (SARS CoV-2) disease (COVID-19) across the U.S., there is a high unmet need for data to inform decision-making regarding social distancing and universal masking. We examined the association of community-level social distancing measures and individual masking with risk of predicted COVID-19 in a large prospective U.S. cohort study of 198,077 participants. Individuals living in communities with the greatest social distancing had a 31% lower risk of predicted COVID-19 compared with those living in communities with poor social distancing. Self-reported masking was associated with a 63% reduced risk of predicted COVID-19 even among individuals living in a community with poor social distancing. These findings provide support for the efficacy of mask-wearing even in settings of poor social distancing in reducing COVID-19 transmission. In the current environment of relaxed social distancing mandates and practices, universal masking may be particularly important in mitigating risk of infection.

## Introduction

In March 2020, most U.S. states implemented community social distancing interventions, including shelter-in-place orders, school closures, bans on large gatherings, and suspension of non-essential businesses, in an attempt to limit transmission of the novel severe acute respiratory syndrome coronavirus 2 (SARS-CoV-2), the cause of coronavirus disease 2019 (COVID-19).^1^ In some areas, these measures appear to have successfully reduced the pace and severity of COVID-19 burden during the initial wave of infections^2,3^, thereby “flattening the curve.” However, lockdowns are not viable as a long-term solution ^4,5^. In addition, despite growing evidence showing that masking can reduce disease transmission^6,7^, adherence to masking recommendations by public health authorities have been variable across the U.S. Given the continued rising burden of COVID-19 across many U.S. communities, there is a high unmet need for data to inform decision-making regarding universal masking in settings in which social distancing is not widely observed.

To date, most evidence on the efficacy of social distancing and universal masking is based on modeling using community-level data in relation to disease burden as assessed through testing, hospitalizations, or mortality^7–9^. Such studies are unable to concurrently account for personal risk factors for infection or optimally assess the latency between social distancing or masking interventions and infection rates given the significant lag between the onset of symptoms, testing, and medical care. Here, we conducted a prospective study in the U.S using a smartphone-based application that collected self-reported, individual-level information on COVID-19 like symptoms, masking and other personal risk factors, in combination with community-level social distancing measures to investigate the relative effectiveness of social distancing and masking policies with the risk of COVID-19.

## Methods

### Study Population

Our study population includes all participants enrolled in the COVID Symptom Study smartphone application (“app”) from March 29, 2020 to July 16, 2020 in the U.S. The app is a freely available program developed by Zoe Global Ltd. in collaboration with researchers and clinicians at Massachusetts General Hospital and King’s College London. Participants using this app reported demographic information and comorbidities at baseline and were encouraged to report on their current health condition daily to allow for the longitudinal, prospective collection of symptoms and COVID-19 testing results^10^. Participants were recruited through general and social media outreach, as well as direct invitations from the investigators of long-running prospective cohorts to study participants^11^. At enrollment, participants provided informed consent to the use of aggregated information for research purposes and agreed to applicable privacy policies and terms of use. This research study was approved by the Partners Human Research Committee (Institutional Review Board Protocol 2020P000909). This protocol is registered with ClinicalTrials.gov (NCT04331509).

### Assessment of predicted COVID-19 and personal risk factors

The information collected through the app has been provided in detail previously^10^. Upon first use, participants were asked to provide baseline demographic factors, including their zip code of residence, and answered separate questions about a suspected risk factors for COVID-19 (Table 1). On first use and upon daily reminders, participants were asked if they felt physically normal, and if not, their symptoms, including fever, persistent cough, fatigue, loss of smell/taste, and diarrhea, among others^10^. Participants were also asked if they had been tested for COVID-19, and if yes, the results (none, negative, waiting, or positive). Beginning on June 12, 2020, participants were also asked if they had worn a face mask when outside the house in the last week (never, sometimes, most of the time, or always). Population density was calculated from Census data for all Zip Code Tabulation Areas (ZCTA) in the U.S. The daily estimated effective reproductive number (Rt), the average number of secondary cases arising from a single case for a given day in each state, was extracted from rt.live^12^. The method and adaptation for estimation of Rt was previously described^13,14^. Because a report of a positive COVID-19 test depends on access to testing and incorporates a variable delay between symptoms and testing, we used a previously published symptom-based classifier that predicts COVID-19 as our primary outcome measure^15^. Between March 24 and April 21, 2020, 2,450,569 UK and 168,293 U.S. individuals reported symptoms and 6,452 UK and 726 U.S. individuals reported a positive COVID-19 test. To build a prediction model, the UK participants were randomly divided into a training set and a test set (ratio: 80:20). Based on the training set, a logistic model generated to predict symptomatic COVID-19 was: Log odds (Predicted COVID-19) = −1.32 - (0.01 x age) + (0.44 x male sex) + (1.75 x loss of smell or taste) + (0.31 x severe or significant persistent cough) + (0.49 x severe fatigue) + (0.39 x skipped meals). The prediction model achieved a sensitivity of 0.65 (95% CI 0.62-0.67) and specificity of 0.78 (95% CI 0.76-0.80) in the test set. In additional validation in the U.S. participants, the prediction model achieved a sensitivity of 0.66 (95% CI 0.62-0.69) and specificity of 0.83 (95% CI 0.82-0.85). To examine the influence of COVID-19 incidence on our results, we included the daily county-level test positive COVID-19 incidence estimated by the Center for Systems Science and Engineering at Johns Hopkins University as a covariate^16,17^.

**Table 1.**
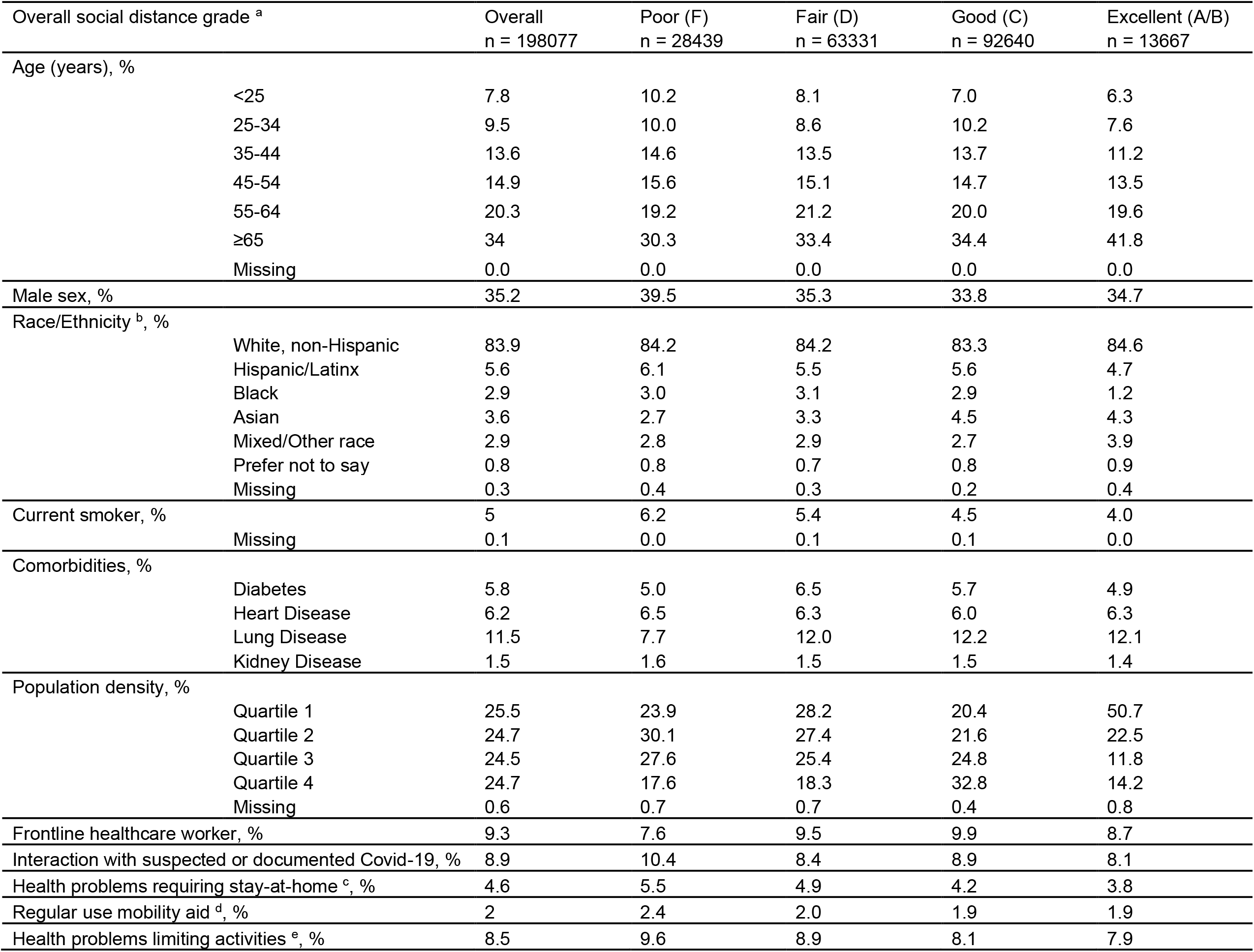
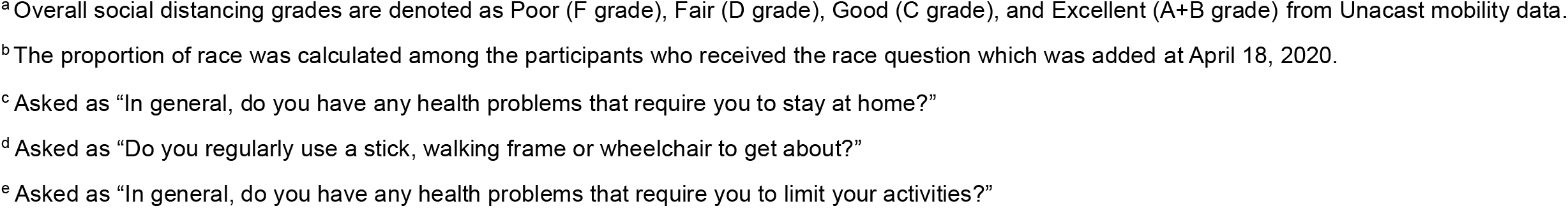
Baseline characteristics of study participants according to overall social distancing grade.

| Overall social distance grade <sup>a</sup> |  | Overall<br>n = 198077 | Poor (F)<br>n = 28439 | Fair (D)<br>n = 63331 | Good (C)<br>n = 92640 | Excellent (A/B)<br>n = 13667 |
| --- | --- | --- | --- | --- | --- | --- |
| Age (years), % |  |  |  |  |  |  |
|  | <25 | 7.8 | 10.2 | 8.1 | 7.0 | 6.3 |
|  | 25-34 | 9.5 | 10.0 | 8.6 | 10.2 | 7.6 |
|  | 35-44 | 13.6 | 14.6 | 13.5 | 13.7 | 11.2 |
|  | 45-54 | 14.9 | 15.6 | 15.1 | 14.7 | 13.5 |
|  | 55-64 | 20.3 | 19.2 | 21.2 | 20.0 | 19.6 |
|  | ≥65 | 34 | 30.3 | 33.4 | 34.4 | 41.8 |
|  | Missing | 0.0 | 0.0 | 0.0 | 0.0 | 0.0 |
| Male sex, % |  | 35.2 | 39.5 | 35.3 | 33.8 | 34.7 |
| Race/Ethnicity <sup>b</sup>, % |  |  |  |  |  |  |
|  | White, non-Hispanic | 83.9 | 84.2 | 84.2 | 83.3 | 84.6 |
|  | Hispanic/Latinx | 5.6 | 6.1 | 5.5 | 5.6 | 4.7 |
|  | Black | 2.9 | 3.0 | 3.1 | 2.9 | 1.2 |
|  | Asian | 3.6 | 2.7 | 3.3 | 4.5 | 4.3 |
|  | Mixed/Other race | 2.9 | 2.8 | 2.9 | 2.7 | 3.9 |
|  | Prefer not to say | 0.8 | 0.8 | 0.7 | 0.8 | 0.9 |
|  | Missing | 0.3 | 0.4 | 0.3 | 0.2 | 0.4 |
| Current smoker, % |  | 5 | 6.2 | 5.4 | 4.5 | 4.0 |
|  | Missing | 0.1 | 0.0 | 0.1 | 0.1 | 0.0 |
| Comorbidities, % |  |  |  |  |  |  |
|  | Diabetes | 5.8 | 5.0 | 6.5 | 5.7 | 4.9 |
|  | Heart Disease | 6.2 | 6.5 | 6.3 | 6.0 | 6.3 |
|  | Lung Disease | 11.5 | 7.7 | 12.0 | 12.2 | 12.1 |
|  | Kidney Disease | 1.5 | 1.6 | 1.5 | 1.5 | 1.4 |
| Population density, % |  |  |  |  |  |  |
|  | Quartile 1 | 25.5 | 23.9 | 28.2 | 20.4 | 50.7 |
|  | Quartile 2 | 24.7 | 30.1 | 27.4 | 21.6 | 22.5 |
|  | Quartile 3 | 24.5 | 27.6 | 25.4 | 24.8 | 11.8 |
|  | Quartile 4 | 24.7 | 17.6 | 18.3 | 32.8 | 14.2 |
|  | Missing | 0.6 | 0.7 | 0.7 | 0.4 | 0.8 |
| Frontline healthcare worker, % |  | 9.3 | 7.6 | 9.5 | 9.9 | 8.7 |
| Interaction with suspected or documented Covid-19, % |  | 8.9 | 10.4 | 8.4 | 8.9 | 8.1 |
| Health problems requiring stay-at-home <sup>c</sup>, % |  | 4.6 | 5.5 | 4.9 | 4.2 | 3.8 |
| Regular use mobility aid <sup>d</sup>, % |  | 2 | 2.4 | 2.0 | 1.9 | 1.9 |
| Health problems limiting activities <sup>e</sup>, % |  | 8.5 | 9.6 | 8.9 | 8.1 | 7.9 |

<sup>a</sup> Overall social distancing grades are denoted as Poor (F grade), Fair (D grade), Good (C grade), and Excellent (A+B grade) from Unacast mobility data.
<sup>b</sup> The proportion of race was calculated among the participants who received the race question which was added at April 18, 2020.
<sup>c</sup> Asked as “In general, do you have any health problems that require you to stay at home?”
<sup>d</sup> Asked as “Do you regularly use a stick, walking frame or wheelchair to get about?”
<sup>e</sup> Asked as “In general, do you have any health problems that require you to limit your activities?”

### Assessment of community social distancing and personal face mask use

We assigned each individual participant a social distancing grade within their communities based on their zip code of residence. We used data provided by Unacast^18^ that estimated county-level social distancing for each calendar day according to the GPS activity of all devices assigned to their longest recorded location. Compared to the same day of the week during the pre-COVID-19 period (defined by Unacast as the four weeks prior to March 8, 2020), Unacast estimated, for each day, the percent reduction in three metrics – metric 1) average distance traveled per device; metric 2) non-essential visitation (e.g., restaurants, department stores, hair salons); and metric 3) human encounters calculated as two devices in close proximity (i.e., spatial distance of ≤50 m and temporal distance of ≤60 minutes)^18^. Unacast assigned grades (A, B, C, D, and F) using predefined cutoff points for each metric and calculated an overall social distancing grade (Supplementary Methods), with grade A indicating the greatest social distancing and F the poorest social distancing. For all analyses, we combined grades A and B due to a limited number of individuals living in counties assigned to grade A. For personal face mask use, we used the individual-level information collected through the app. Beginning on June 12, app users received supplementary questions regarding use of face masks based on the query “In the last week, did you wear a face mask when outside the house?”.

### Statistical Analysis

We conducted prospective analyses after excluding participants who had any symptom related to COVID-19 or who had tested positive for COVID-19 prior to start of follow-up to minimize collider bias. Follow-up time started when participants first reported on the app and accrued until they developed predicted COVID-19, or the time of last data entry prior to July 16^th^, whichever occurred first. We used updated, time-varying community social distancing exposure data as our primary independent variable. Community-level social distancing exposure data and corresponding follow-up time was mapped to each individual and updated each time they logged in the app to provide updated symptom information. We also used time-varying masking exposure data for the association between self-reported personal use of masks and predicted COVID-19. Cox proportional hazards regression models stratified by age, state, and calendar date at study entry were used to calculate unadjusted and multivariable adjusted hazard ratios (HRs) and 95% confidence intervals (CIs) of predicted COVID-19. Covariates were selected *a priori* based on putative risk factors and included race (white, Black, Asian, other race), sex (male, female), population density (quartiles), current smoking, work as a frontline healthcare worker, interaction with suspected or documented COVID-19, and history of diabetes, heart disease, lung disease, and kidney disease (each yes/no). Missing data for categorical variables was included as a missing indicator.

To minimize any variation of estimated daily social distancing grade associated with day of the week (e.g. Sunday vs. Monday), we used a seven-day average of community social distancing grade as the exposure for each participant. We first examined the latency between community social distancing grade and predicted COVID-19 using varying lag times (0 days, 7 days, 14 days, 21 days, and 28 days). For example, for a latency of 7 days, we used social distancing grade exposure on April 1 for predicted COVID-19 outcome measures on April 8, grade on April 2 for follow-up on April 9 and so forth (Supplemental Figure 1). For subgroup analysis according to daily state-level Rt, we used a 21-day latency since this corresponded to the start of the seven-day average social distancing exposure with a 14-day latency. Two-sided p-values of <0.05 were considered statistically significant for main analyses. All statistical analyses were performed using R software, version 3.6.1 (R Foundation).

## Results

Between March 29 and July 16, 2020, we enrolled 277,798 participants who provided baseline information. We excluded 79,721 individuals who did not live in a county with available Unacast data, reported any symptoms or a positive COVID 19 test at enrollment, had <24 hours of follow-up time or who reported a positive COVID-19 test or symptoms of predicted COVID-19 within 24 hours of enrollment. This left 198,077 participants in our prospective inception cohort, in which we subsequently documented 4,488 cases of predicted COVID-19 over 11,403,773 person-days of follow-up. Compared to others, individuals who lived in communities with poor social distancing (Grade=F) at baseline were younger, more likely to be male, more likely to smoke currently, have less lung disease, and had more interaction with suspected or documented COVID-19 individuals (Table 1). In contrast, individuals living in communities with excellent social distancing (Grade=A/B) were older and more likely to live in areas with lower population density (Table 1).

### Risk of predicted COVID-19 according to overall community social distancing grade at various time lags

To test the association between community-level social distancing and risk of subsequent predicted COVID-19, we evaluated lag times of 7 to 28 days. Living in a community with greater social distancing grade (F to A/B) was associated with a lower risk of predicted COVID-19 for all lag times evaluated (Table 2). The maximal association was first observed with a fourteen-day lag and the benefit plateaued beyond that time period (Figure 1). Compared to participants living in communities with overall poor social distancing (Grade=F), the multivariable HRs for predicted COVID-19 at 14 days were 0.85 (95% CI 0.77-0.95) for fair (Grade=D), 0.80 (95% CI 0.70-0.91) for good (Grade=C), and 0.69 (95% CI 0.55-0.86) for excellent (Grade=A/B) social distancing (*P*_linear-trend_ <.001) after adjusting for personal risk factors for COVID-19 (Table 2). There was a negative but not statistically significant association with a 0-day lag. When we further adjusted for county-level test positive COVID-19 incidence in the community at the time of assessment for the social-distancing measures, we observed similar results (multivariable HR, 0.67; 95% CI 0.53-0.85) for excellent social distancing (Grade=A/B) compared to participants living in communities with overall poor social distancing (Grade=F). For subsequent analyses, we focused on models using a fourteen-day latency since the reduction in predicted COVID-19 appeared maximal at 14 days, and this is considered a plausible interval for exposure to symptom-based disease prediction.

**Table 2.** Risk of predicted Covid-19 according to living in a community with overall social distancing grade at various time lags.

| Overall social distance grade <sup>a</sup> | Poor (F) | Fair (D) | Good (C) | Excellent (A/B) | P value for Trend <sup>b</sup> |
| --- | --- | --- | --- | --- | --- |
| Day -0 |  |  |  |  |  |
| No. of Case/ Person-time (days) | 1854/6036928 | 1321/3388613 | 1164/1790701 | 149/187531 |  |
| Model 1 HR (95% CI) <sup>c</sup> | 1 [Reference] | 0.92 (0.83-1.02) | 0.89 (0.78-1.01) | 0.86 (0.69-1.06) | 0.06 |
| Model 2 HR (95% CI) <sup>d</sup> | 1 [Reference] | 0.93 (0.84-1.02) | 0.89 (0.78-1.01) | 0.84 (0.68-1.05) | 0.06 |
| Day -7 |  |  |  |  |  |
| No. of Case/ Person-time (days) | 1631/5328173 | 1373/3526016 | 1334/2283244 | 150/266340 |  |
| Model 1 HR (95% CI) | 1 [Reference] | 0.90 (0.81-1.00) | 0.86 (0.76-0.98) | 0.77 (0.62-0.96) | 0.01 |
| Model 2 HR (95% CI) | 1 [Reference] | 0.89 (0.80-0.99) | 0.85 (0.75-0.97) | 0.78 (0.63-0.97) | 0.01 |
| Day -14 |  |  |  |  |  |
| No. of Case/ Person-time (days) | 1538/4650075 | 1457/3680269 | 1352/2733816 | 141/339613 |  |
| Model 1 HR (95% CI) | 1 [Reference] | 0.85 (0.77-0.95) | 0.79 (0.70-0.90) | 0.68 (0.54-0.84) | <.001 |
| Model 2 HR (95% CI) | 1 [Reference] | 0.85 (0.77-0.95) | 0.80 (0.70-0.91) | 0.69 (0.55-0.86) | <.001 |
| Day -21 |  |  |  |  |  |
| No. of Case/ Person-time (days) | 1651/4106239 | 1441/3843175 | 1256/3060696 | 140/393663 |  |
| Model 1 HR (95% CI) | 1 [Reference] | 0.82 (0.74-0.91) | 0.74 (0.65-0.84) | 0.68 (0.55-0.85) | <.001 |
| Model 2 HR (95% CI) | 1 [Reference] | 0.82 (0.74-0.91) | 0.74 (0.65-0.84) | 0.69 (0.56-0.86) | <.001 |
| Day - 28 |  |  |  |  |  |
| No. of Case/ Person-time (days) | 1796/3731565 | 1389/3987114 | 1168/3261867 | 135/423186 |  |
| Model 1 HR (95% CI) | 1 [Reference] | 0.82 (0.74-0.90) | 0.75 (0.66-0.86) | 0.69 (0.55-0.86) | <.001 |
| Model 2 HR (95% CI) | 1 [Reference] | 0.82 (0.74-0.91) | 0.75 (0.66-0.86) | 0.70 (0.55-0.87) | <.001 |

Abbreviations: HR (hazard ratio), CI (confidence interval)
<sup>a</sup> Overall social distancing grades are denoted as Poor (F grade), Fair (D grade), Good (C grade), and Excellent (A+B grade). Overall social grade categories (A, B, C, D, and F) are provided by Unacast.
<sup>b</sup> P value for trend is calculated using the median value of each category as a continuous variable.
<sup>c</sup> Model 1 was stratified by age (<25, 25-34, 35-44, 45-54, 55-64, or ≥65), state, and calendar date at study entry.
<sup>d</sup> Model 2 was stratified by age (<25, 25-34, 35-44, 45-54, 55-64, ≥65), state, and calendar date at study entry and further adjusted for race (white, black, Asian, or other), sex (male or female), population density of residence (quartiles), current smoking, frontline healthcare worker, interaction with suspected or documented Covid-19, history of diabetes, heart disease, lung disease, and kidney disease (each yes or no).

**Figure 1.**
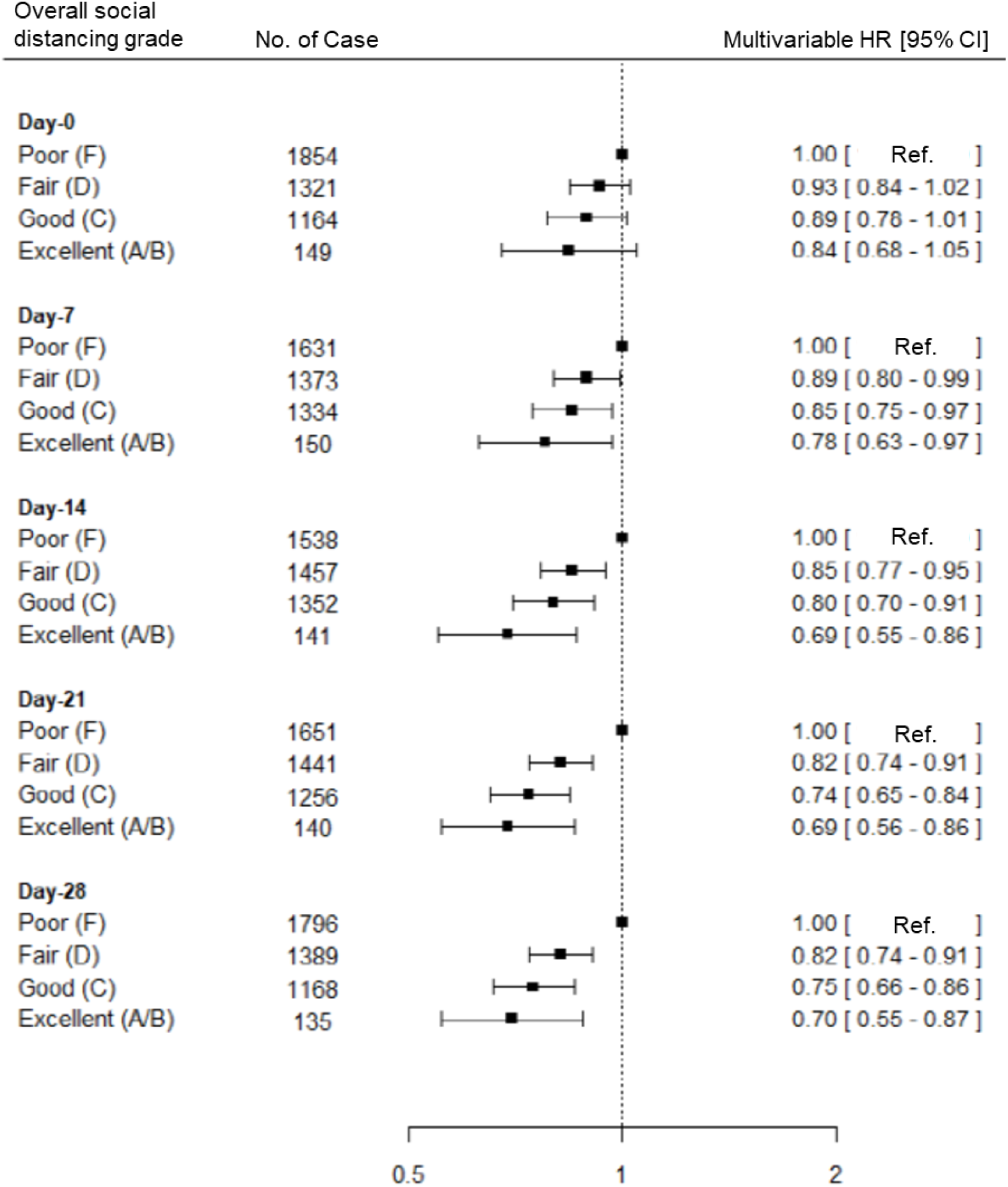
Risk of predicted Covid-19 according to living in a community with overall social distancing grade ^a^ at various time lags. Abbreviations: HR (hazard ratio), CI (confidence interval) ^a^ Overall social distancing grades are denoted as Poor (F grade), Fair (D grade), Good (C grade), and Excellent (A+B grade). Overall social grade categories (A, B, C, D, and F) are provided by Unacast. Multivariable model was stratified by age (<25, 25-34, 35-44, 45-54, 55-64, ≥65), state, and calendar date at study entry and further adjusted for race (white, black, Asian, or other), sex (male or female), population density of residence (quartiles), current smoking, frontline healthcare worker, interaction with suspected or documented Covid-19, history of diabetes, heart disease, lung disease, and kidney disease (each yes or no).

### Risk of predicted COVID-19 according to community social distancing metrics and demographics

We also assessed the three individual components of the Unacast social distancing grade: including average distance traveled, non-essential visitation, and human encounters (Table 3). Reduction in average distance traveled (HR, 0.78; 95% CI 0.65-0.92 <25% versus >55%) and non-essential visitation (HR, 0.79; 95% CI 0.70-0.89 <55% versus >65%) were both associated with lower risk of predicted COVID-19. The reduction in human encounters, based on phone-to-phone proximity measures, was not associated with lower risk of predicted Covid-19. In subgroup analyses, the association of social distancing grade and COVID-19 appeared to differ according to age (*P*_interaction_=0.001). The benefit of increasing social distancing from Poor (F) to Excellent (A/B) was greatest among the middle-age participants (35-55 years, HR, 0.47; 95% CI 0.26-0.86), than among younger (age <35 years) or older participants (>55). We assessed for effect modification by other demographic including race, sex, and health problems limiting activities, and found no significant interactions between social distancing grades and these factors (all *P*_interaction_ >0.05; Supplemental Table 2).

**Table 3.** Risk of predicted Covid-19 within 14 days according to individual metrics of social distancing ^a^.

| Social distance grade <sup>b</sup> | Poor (F) | Fair (D) | Good (C) | Excellent (A/B) | P value for Trend <sup>c</sup> |
| --- | --- | --- | --- | --- | --- |
| Metric 1: Percent reduction in average distance traveled |  |  |  |  |  |
|  | < 25% | 25-40% | 40-55% | >55% |  |
| No. of Case/ Person-time (days) | 1421/4157983 | 1233/3286302 | 1352/2994955 | 482/964532 |  |
| Model 1 HR (95% CI) <sup>d</sup> | 1 [Reference] | 0.84 (0.76-0.93) | 0.78 (0.69-0.88) | 0.82 (0.70-0.96) | <.001 |
| Model 2 HR (95% CI) <sup>e</sup> | 1 [Reference] | 0.84 (0.75-0.93) | 0.77 (0.68-0.88) | 0.78 (0.65-0.92) | <.001 |
| Metric 2: Percent reduction in non-essential visitation |  |  |  |  |  |
|  | <55% | 55-60% | 60-65% | >65% |  |
| No. of Case/ Person-time (days) | 2164/6338342 | 445/1149430 | 533/1171405 | 1255/2480483 |  |
| Model 1 HR (95% CI) | 1 [Reference] | 0.84 (0.75-0.95) | 0.85 (0.75-0.96) | 0.79 (0.71-0.88) | <.001 |
| Model 2 HR (95% CI) | 1 [Reference] | 0.84 (0.75-0.95) | 0.85 (0.76-0.97) | 0.79 (0.70-0.89) | <.001 |
| Metric 3: Percent reduction in human encounters |  |  |  |  |  |
|  | <40% | 40-74% | 74-82% | >82% |  |
| No. of Case/ Person-time (days) | 3409/8622754 | 441/1099267 | 153/417628 | 485/1264123 |  |
| Model 1 HR (95% CI) | 1 [Reference] | 1.01 (0.90-1.12) | 0.99 (0.83-1.18) | 0.96 (0.86-1.06) | 0.60 |
| Model 2 HR (95% CI) | 1 [Reference] | 1.02 (0.91-1.14) | 1.00 (0.84-1.20) | 0.95 (0.84-1.08) | 0.78 |
| Overall social distancing grade <sup>f</sup> |  |  |  |  |  |
| No. of Case/ Person-time (days) | 1538/4650075 | 1457/3680269 | 1352/2733816 | 141/339613 |  |
| Model 1 HR (95% CI) | 1 [Reference] | 0.85 (0.77-0.95) | 0.79 (0.70-0.90) | 0.68 (0.54-0.84) | <.001 |
| Model 2 HR (95% CI) | 1 [Reference] | 0.85 (0.77-0.95) | 0.80 (0.70-0.91) | 0.69 (0.55-0.86) | <.001 |

Abbreviations: HR (hazard ratio), CI (confidence interval)
<sup>a</sup> Day-14 is applied for models.
<sup>b</sup> Social distancing grades are denoted as Poor (F grade), Fair (D grade), Good (C grade), and Excellent (A+B grade). The cut-offs for Metric 1,2, and 3 and overall social grade categories (A, B, C, D, and F) are provided by Unacast.
<sup>c</sup> P value for trend is calculated using the median value of each category as a continuous variable.
<sup>d</sup> Model 1 was stratified by age (<25, 25-34, 35-44, 45-54, 55-64, or ≥65), state, and calendar date at study entry.

Furthermore, to evaluate whether the impact of social distancing on risk of predicted COVID-19 was modified by local transmissibility, we performed subgroup analysis according to Rt. During epidemic slowing/maintenance period (Rt ≤1.0), compared to participants living in communities with overall poor social distancing (Grade=F), the multivariable HRs for predicted COVID-19 were 0.88 (95% CI 0.75-1.02) for fair (Grade=D), 0.79 (95% CI 0.66-0.96) for good (Grade=C), and 0.64 (95% CI 0.48-0.87) for excellent (Grade=A/B) social distancing (*P*_linear-trend_ =0.004) after adjusting for personal risk factors for COVID-19 (Supplemental Table 5). This trend was also observed with similar magnitudes albeit with borderline significance (*P*_linear-trend_ =0.08) during the epidemic growth period (Rt >1.0).

### Risk of predicted COVID-19 according to personal face mask use

We examined the association between self-reported personal use of a face mask and risk of predicted COVID-19 among the 139,690 participants who provided this information. Compared to individuals who reported never using a face mask, individuals who reported using (sometimes, most of the time, or always) a face mask had a multivariable HR for predicted COVID-19 of 0.35 (95% CI 0.30-0.42; Table 4).

**Table 4.**
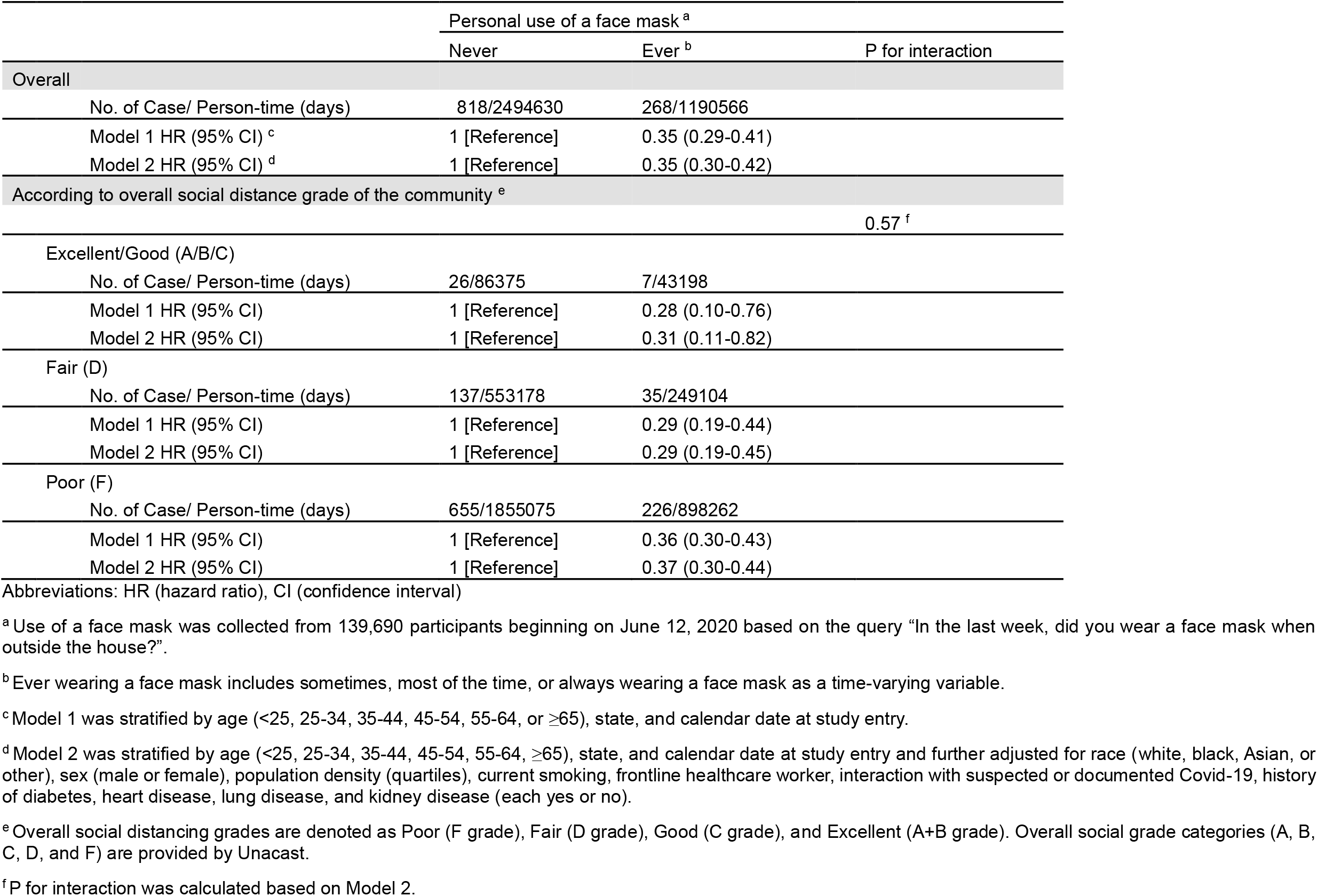
Personal use of a face mask outside the home and risk of predicted Covid-19.

Ever using a face mask was associated with reduced predicted COVID-19, with adjusted HRs of 0.31 (95% CI 0.11-0.82) among individuals living in communities with excellent or good social distancing grade, 0.29 (95% CI 0.19-0.45) for those living in communities with fair social distancing grade, and 0.37 (95% CI, 0.30–0.44) for those living in communities with poor social distancing grade (*P*_interaction_=0.57). The results remained similar after additional adjustment for actual COVID-19 incidence. Furthermore, observed associations were not substantially different when analyses were restricted to participants living in Texas, Arizona, California, and Florida (HR, 0.35; 95% CI 0.26-0.47), states which were among the states in which social distancing policy was relaxed earlier during the initial phase of the pandemic. Finally, the association of personal use of a face mask and predicted COVID-19 did not appear to substantially different according to *Rt* (Supplemental Table 6).

## Discussion

In this prospective study of 198,077 participants using a real-time mobile phone application in U.S., we observed that individuals living in communities with the greatest social distancing had a 31% lower risk of predicted COVID-19 compared with those living in communities with poor social distancing, with maximum benefit evident after a latency period of 14 days. Furthermore, among individuals living in communities with poor social distancing, personal use of a face mask outside of the home at least sometimes was associated with a 63% reduced risk of predicted COVID-19 compared to individuals who never wore a face mask.

Notably, a reduction in average distance traveled and non-essential visitation in the community was protective for predicted COVID-19. In contrast, close contact as measured by human encounters was not associated with predicted COVID-19. This suggests that average distance traveled and non-essential visitation, as measures of independent mobility, may be more reflective of effective social distancing than measures based on assessing proximity between two devices. It is also possible that the criterion to define human encounters based on devices <50 meters apart may not be optimal to study COVID-19 transmission. In subgroup analysis, we did not observe the inverse associations between living in communities with the greater social distancing and risk of COVID-19 among individuals aged greater than 55 years, having health problems requiring stay-at-home, and regularly using mobility aids. For those individuals, living in a community with the greatest social distancing may not play an important role in reducing COVID-19 risk due to their limited mobility and lower likelihood of social interaction in crowded spaces. Noticeably, the inverse association between living in a community with greater social distancing and risk of predicted COVID-19 was most consistently observed among younger individuals without significant health problems or limitations in mobility.

We observed that the disease burden of COVID-19 at the start of the social distancing measurement did not influence the association of social distancing and personal use of a face mask with risk of predicted COVID-19. We also observed that protective effect of social distancing on predicted COVID-19 was present both in areas where the epidemic was slowing or maintained (Rt ≤1.0) as well as in areas where COVID-19 was actively spreading (Rt>1.0). We similarly observed that the benefit of personal use of a face mask was observed in regions and time periods in which there was epidemic slowing/maintenance or growth. These findings imply that baseline risk did not impact the relative benefits of social distancing policies and/or face mask use, although it is remains possible that the absolute reduction in risk is greater in areas with higher burden of COVID-19.

In our study, we used predicted COVID-19 as a proxy for a positive COVID-19 test due to the small number of COVID-19 test positive app users during the study period. The small fraction of positive COVID-19 tests among all participants (0.17%) may be largely influenced by the limited availability of COVID-19 testing during the study period. A recent study demonstrated that more than 80% of individuals with a COVID-19 infection in the U.S. went undetected in March 2020^19^. Moreover, another study in 10 sites across U.S. reported that the estimated number of COVID-19 infections was 6 to 24 times greater per site than the number of reported from March 23 to May 12^20^. Nevertheless, despite the limited power, we found a protective but not statistically significant association between community social distancing and risk of a positive COVID-19 test (Supplemental table 3). Therefore, this association between the social distancing observed within one’s community and a positive COVID-19 test should be further investigated in studies in which there was a higher prevalence of testing.

Our findings are consistent with previous ecological studies investigating the effect of social distancing on risk of COVID-19^21–28^. In one recent study that also used estimates of social distancing based on Unacast data, each one-unit increase in social distancing was associated with a 29% reduced risk of COVID-19 incidence and a 35% reduced risk of COVID-19 mortality^22^ at the county-level. In a separate study, COVID-19 epidemic case growth rates declined by approximately 1% per day beginning four days after statewide social distancing measures were implemented^21^. In addition, estimated rates of COVID-19 cases were increased in border counties in Iowa which did not issue a stay-at-home order compared with border counties in Illinois which did issue a stay-at-home order^23^. Another study based on 149 countries demonstrated that any physical distancing intervention was associated with 13% reduced risk of COVID-19 incidence^3^. These finding add to this body of evidence as we estimate the impact of social distancing in the community on individual-level outcomes.

Other studies have shown that masking is associated with a lower risk of COVID-19 on a population-scale^6,7,13,25,29^. In one recent study among health care workers, universal masking in a hospital setting was associated with a lower rate of COVID-19^6,30^. A recent meta-analysis demonstrated that face mask use was associated with a 85% reduced risk of viral infection causing COVID-19, SARS (severe acute respiratory syndrome), or MERS (Middle East respiratory syndrome) in both health care and non-health care settings^7^. While the role of a face mask in protecting other individuals is well-recognized, we observed that a face mask may also protect individuals who wear them, as has been described by others^29^. Alternatively, participants who generally are willing to wear a face (or self-report such) mask may also engage in overall healthier behaviors.

This study has several strengths. First, we used a mobile application to rapidly collect prospective data from a large population on known or suspected COVID-19 personal risk factors, such as mask wearing. This is a significant advantage over existing studies which cannot concurrently examine the impact of personal interventions to reduce exposure risk with community-scale data. Second, we collected data from participants initially free of a positive COVID-19 test and any symptoms, which allowed a prospective assessment of incident symptoms with minimal recall or collider bias. Third, we assessed COVID-19 incidence according to a validated symptom assessment which minimizes the biases associated with geographic variation in access^31^ to COVID-19 testing on estimates of COVID-19 incidence, which may bias effect estimates away from or towards the null (e.g. social distancing associated with reduced test access or increased test seeking behavior). This also allows us to better assess the impact of social distancing on COVID-19 according to different latency periods since it minimizes the time delay between onset of infection, obtaining a test, and reporting of the result, which has been estimated to be delayed by as long as a week in some areas of the U.S.^32,33^.

There are several limitations to our study. First, our information on risk factors and symptoms are collected by self-report. Although information based on clinical records and testing would be more accurate, given the rapid pace of the pandemic and the limited availability of medical care and testing, self-reported information is more feasible to collect longitudinally and prospectively among a large number of participants and minimizes recall bias or selection bias (e.g. preferentially capturing severe cases through hospitalization records or death reports). Second, since our cohort is not a random sampling of the population, there remains a possibility for selection or collider bias, as well as generalizability. We tried to minimize collider bias in this study by excluding participants who reported having COVID-19 symptoms prior to joining the app. Although this limitation is inherent to any study requiring voluntary provision of information, we acknowledge that data collection through smartphone adoption has comparatively lower penetrance among certain socioeconomic groups and older adults and that participants of an app study may have differential likelihood of reporting symptoms^34^. Third, it is possible that the personal risk factors for COVID-19 that we assessed here, such as wearing a face mask, may be confounded by other behaviors that reduce infection risk, as well as whether users are accurately self-reporting these behaviors. Fourth, the social distancing metrics used as an exposure are not reflective of actual user mobility. There may be non-differential misclassification of exposure status by region, if county-level factors are correlated with the individual-level heterogeneity of each mobility metric (e.g. younger app users in an urban area with high mobility). Fifth, our analysis was focused on symptomatic COVID-19. However, it is likely that an association between social distancing and masking with risk of asymptomatic spread would be similar. Lastly, while personal face mask use and other covariates were based on individual level data reported through the app, the social distancing measures are based on regionally aggregated data assigned to each app user.

In conclusion, within a large population-based sample of individuals in the U.S, we demonstrated a significantly reduced risk of predicted COVID-19 infection among individuals living in communities with a greater social distancing grade at 14 days either in regions or time periods experiencing either epidemic slowing or growth.

Among participants who lived in a community with poor social distancing, wearing a face mask was associated with reduced risk. These findings provide additional support for the efficacy of non-pharmaceutical interventions in reducing COVID-19 incidence and spread and suggest that the benefits of such interventions will become most evident at 14 days after implementation. Encouraging universal masking may be particularly important to limit the continued spread of COVID-19 as social distancing mandates continue to be relaxed.

## Data Availability

Data collected in the app are being shared with other health researchers through the NHS-funded Health Data Research UK (HDRUK)/SAIL consortium, housed in the UK Secure e-Research Platform (UKSeRP) in Swansea. Anonymized data can be shared with bonafide researchers via HDRUK, provided the request is made according to their protocols and is in the public interest (see https://healthdatagateway.org/detail/9b604483-9cdc-41b2-b82c-14ee3dd705f6). Data updates can be found at https://covid.joinzoe.com.

## ACKNOWLEDGEMENTS

We would like to thank to all of the participants who entered data into the app, including study volunteers enrolled in cohorts within the Coronavirus Pandemic Epidemiology (COPE) consortium. We thank the staff of Zoe Global Ltd., the Department of Twin Research at King’s College London, and the Clinical and Translational Epidemiology Unit at Massachusetts General Hospital.

