## Supplementary for "Association of social distancing and masking with risk of COVID-19"

**Table of Contents**

**Supplemental Methods** ... *2*

**Supplemental Figure 1. Latency between community social distancing grade and predicted Covid-19 at various time intervals (7, 14, and 21 days latency)** ... *3*

**Supplemental Figure 2. Overall community social distancing grade in the U.S. over time** ... *4*

**Supplemental Table 1. Baseline characteristics of study participants according to personal use of a face mask** *5*

**Supplemental Table 2. Risk of predicted symptomatic COVID-19 according to overall social distancing grade within subgroups** ... *7*

**Supplemental Table 3. Risk of testing positive COVID-19 according to overall social distancing grade** ... *8*

**Supplemental Table 4. Risk of testing positive COVID-19 according to personal use of a face mask** ... *9*

**Supplemental Table 5. Risk of predicted COVID-19 according to overall social distancing grade within Rt subgroups** ... *10*

**Supplemental Table 6. Risk of predicted COVID-19 according to Personal use of a face mask within Rt subgroups** ... *11*

**References** ... *12*

### Supplemental Methods

#### Community social distancing grade

Community social distancing was graded by Unacast for each metric 1, 2, and 3 and overall grade assigned as A, B, C, D, and F [1]. For metric 1, percent reduction in average distance traveled per device are categorized and graded as A: >70% decrease, B: 55-70% decrease, C: 40-55% decrease, D: 25-40% decrease, and F: <25% decrease or increase. For metric 2, percent reduction in non-essential visitation are categorized and graded as A: >70% decrease, B: 65-70% decrease, C: 60-65% decrease, D: 55-60% decrease, and F: <55% decrease or increase. For metric 3, percent reduction in human encounters are categorized and graded as A: >94% decrease, B: 94-82% decrease, C: 82-74% decrease, D: 74-40% decrease, and F: <40% decrease or increase. The overall grade was calculated based on metric 1, metric 2, and metric 3 as the average between the three numeric grades by Unacast.

For metric 2, non-essential visitation include (but are not limited to): restaurants (multiple kinds), department and clothing stores, jewelers, consumer electronics stores, cinemas and theaters, office supply stores, spas and hair salons, gyms and fitness/recreation facilities, car dealerships, hotels, craft, toy, and hobby shops.

Social distancing data was provided at the county-level. Data are not available for counties with a population less than 1,000; where less than 100 smartphone devices were observed for 70% of the days during the pre-COVID-19 period; or where less than 5 non-essential venues or 100 non-essential venue visits occurred during the pre-COVID-period.

We then assigned the county-level social distancing grade to each participant according to their zip code, for which we used a U.S. Department of Housing and Urban Development Zip Code Crosswalk files to link ZIP Codes to counties.

**Supplemental Figure 1. Latency between community social distancing grade exposure and predicted Covid-19 outcome at various time intervals (7, 14, and 21 days latency)**

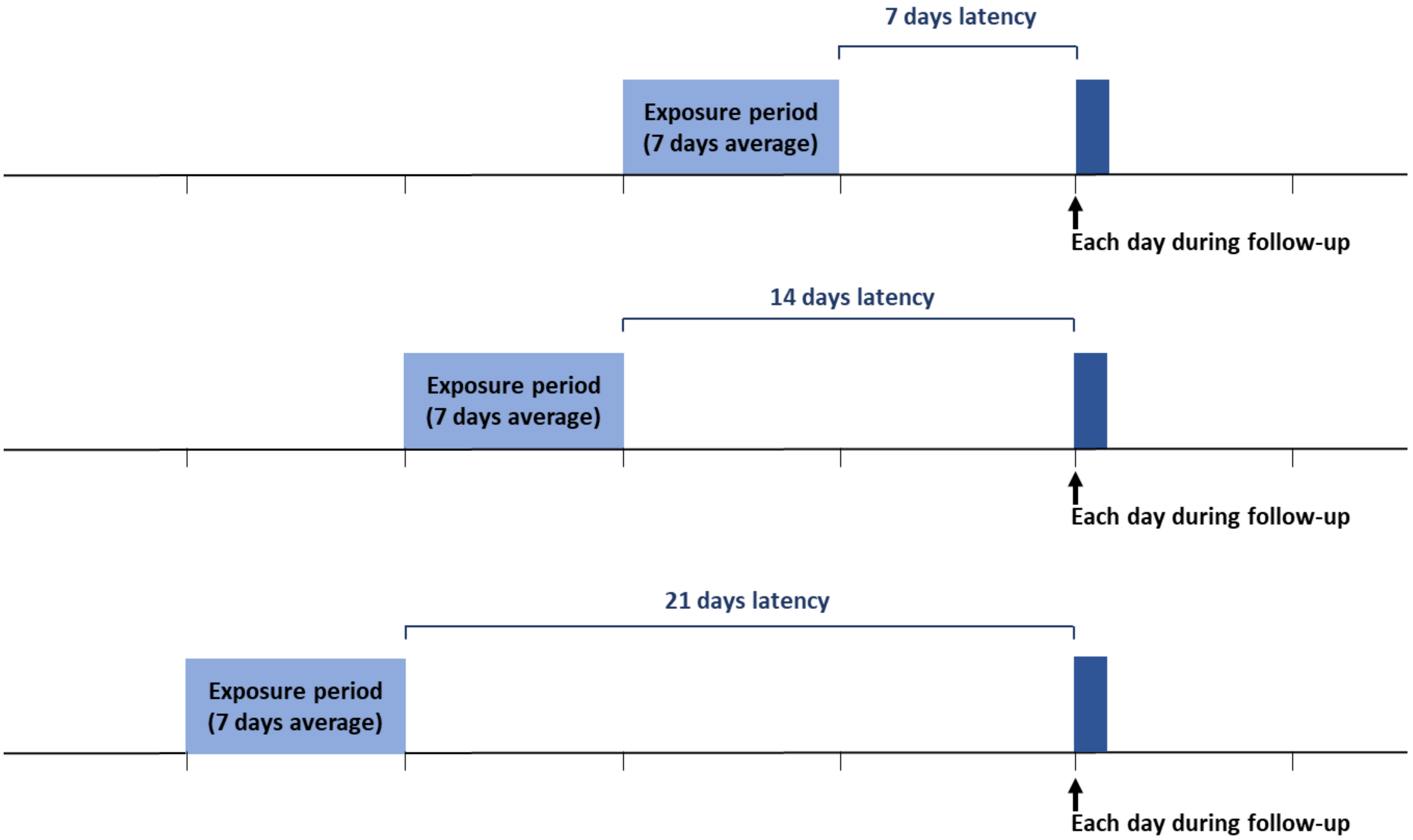

**Supplemental Figure 2. Overall community social distancing grade in the U.S. [a] over time**

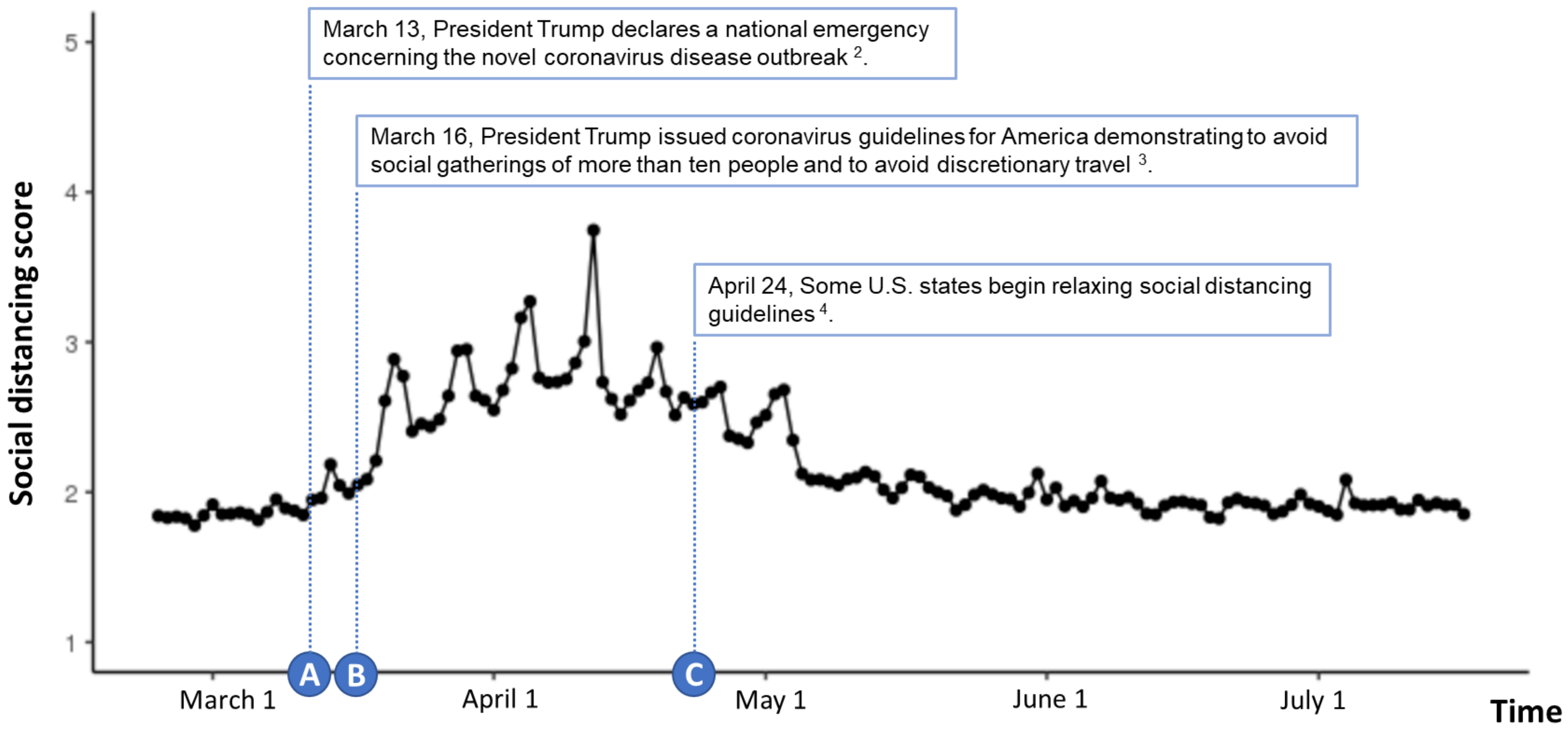


[a] Social distancing score is averaged across the counties in U.S. Excellent (A/B); social distancing score is greater than 4. Good (C); social distancing score is greater than 2.5 and less than 4. Fair (B); social distancing score is greater than 1.5 and less than 2.5. Poor (F); social distancing score is less than 1.5.

**Supplemental Table 1. Baseline characteristics of study participants according to personal use of a face mask**

| Personal use of a face mask [a] | | Overall n = 132857 | Never n = 92024 | Ever [b] n = 40833 |
|---|---|---|---|---|
| Age (years), % | | | | |
| | <25 | 5.8 | 5.4 | 6.7 |
| | 25-34 | 7.0 | 6.2 | 8.7 |
| | 35-44 | 11.3 | 10.2 | 13.7 |
| | 45-54 | 13.6 | 12.8 | 15.4 |
| | 55-64 | 21.4 | 21.8 | 20.6 |
| | ≥65 | 40.8 | 43.5 | 34.8 |
| | Missing | 0.0 | 0.0 | 0.0 |
| Male sex, % | | 35.2 | 35.7 | 34.1 |
| Race/Ethnicity [c], % | | | | |
| | White, non-Hispanic | 82.5 | 83.2 | 80.8 |
| | Hispanic/Latinx | 4.3 | 3.7 | 5.7 |
| | Black | 2.5 | 2.3 | 3.1 |
| | Asian | 3.4 | 3.3 | 3.8 |
| | Mixed/Other race | 2.5 | 2.4 | 2.8 |
| | Prefer not to say | 1.0 | 1.0 | 1.0 |
| | Missing | 3.8 | 4.2 | 2.8 |
| Current smoker, % | | 3.8 | 3.5 | 4.6 |
| | Missing | 0.0 | 0.0 | 0.0 |
| Comorbidities, % | | | | |
| | Diabetes | 5.9 | 6.2 | 5.4 |
| | Heart Disease | 6.9 | 7.2 | 6.4 |
| | Lung Disease | 11.0 | 11.2 | 10.5 |
| | Kidney Disease | 1.6 | 1.7 | 1.5 |
| Population density, % | | | | |
| | Quartile 1 | 24.5 | 25.0 | 23.6 |
| | Quartile 2 | 24.7 | 24.8 | 24.4 |
| | Quartile 3 | 25.0 | 25.0 | 25.2 |
| | Quartile 4 | 25.2 | 24.7 | 26.2 |
| | Missing | 0.6 | 0.5 | 0.6 |
| Frontline healthcare worker, % | | 7.7 | 7.1 | 9.1 |
| Interaction with suspected or documented Covid-19, % | | 7.8 | 7.0 | 9.6 |
| Health problems requiring stay-at-home [d], % | | 4.5 | 4.3 | 4.9 |
| Regular use mobility aid [e], % | | 2.1 | 2.2 | 2.0 |
| Health problems limiting activities [f], % | | 8.6 | 8.4 | 8.9 |

[a] Use of a face mask was collected from 139,690 participants beginning on June 12, 2020 based on the query "In the last week, did you wear a face mask when outside the house?".

[b] Ever wearing a face mask includes sometimes, most of the time, or always wearing a face mask.

[c] The proportion of race was calculated among the participants who received the race question which was added at April 18, 2020.

[d] Asked as "In general, do you have any health problems that require you to stay at home?"

[e] Asked as "Do you regularly use a stick, walking frame or wheelchair to get about?"

[f] Asked as "In general, do you have any health problems that require you to limit your activities?"

**Supplemental Table 2. Risk of predicted COVID-19 according to overall social distancing grade within subgroups [a]**

| Variables | | No. of Case | Multivariable HR (95% CI) [b] | | P value for Interaction |
|---|---|---|---|---|---|
| | | | Poor (F) | Excellent (A/B) | |
| Age, years | | | | | |
| | ≤ 35 | 619 | 1 [Reference] | 0.73 (0.41-1.31) | 0.001 |
| | 35-55 | 688 | 1 [Reference] | 0.47 (0.26-0.86) | |
| | > 55 | 372 | 1 [Reference] | 1.10 (0.61-1.97) | |
| Race | | | | | |
| | White, non-Hispanic | 1228 | 1 [Reference] | 0.62 (0.33-1.16) | 0.41 |
| | Black/Hispanic | 170 | 1 [Reference] | 0.83 (0.35-1.96) | |
| | Asian/Others | 113 | 1 [Reference] | 0.96 (0.38-2.45) | |
| Sex | | | | | |
| | Male | 801 | 1 [Reference] | 0.62 (0.34-1.12) | 0.52 |
| | Female | 878 | 1 [Reference] | 0.55 (0.30-1.00) | |
| Population density | | | | | |
| | ≤ median | 806 | 1 [Reference] | 0.50 (0.28-0.89) | 0.004 |
| | > median | 860 | 1 [Reference] | 0.91 (0.48-1.74) | |
| Interaction with suspected or documented Covid-19 | | | | | |
| | Yes | 272 | 1 [Reference] | 0.52 (0.25-1.08) | 0.63 |
| | No | 1405 | 1 [Reference] | 0.59 (0.33-1.05) | |
| Health problems requiring stay-at-home [c] | | | | | |
| | Yes | 138 | 1 [Reference] | 1.21 (0.56-2.63) | 0.02 |
| | No | 1541 | 1 [Reference] | 0.55 (0.31-0.97) | |
| Regular use mobility aid [d] | | | | | |
| | Yes | 48 | 1 [Reference] | 2.18 (0.85-5.62) | 0.002 |
| | No | 1631 | 1 [Reference] | 0.56 (0.31-0.99) | |
| Health problems limiting activities [e] | | | | | |
| | Yes | 250 | 1 [Reference] | 0.72 (0.36-1.47) | 0.27 |
| | No | 1429 | 1 [Reference] | 0.54 (0.30-0.97) | |

Abbreviations: HR (hazard ratio), CI (confidence interval)

[a] Day-14 is applied for models.

[b] Multivariable models are adjusted for the same covariates as the model 2 in Table 2.

[c] Asked as “In general, do you have any health problems that require you to stay at home?”

[d] Asked as “Do you regularly use a stick, walking frame or wheelchair to get about?”

[e] Asked as “In general, do you have any health problems that require you to limit your activities?”

**Supplemental Table 3. Risk of testing positive COVID-19 according to overall social distancing grade**

| Overall social distance grade | Poor (F) | Fair (D) | Good (C) | Excellent (A/B) | P value for Trend |
|---|---|---|---|---|---|
| No. of Case/ Person-time (days) | 83/5043451 | 90/3973673 | 163/2968330 | 8/365041 | |
| Model 1 HR (95% CI) | 1 [Reference] | 0.67 (0.43-1.05) | 0.93 (0.57-1.52) | 0.49 (0.21-1.16) | 0.90 |
| Model 2 HR (95% CI) | 1 [Reference] | 0.70 (0.44-1.10) | 0.91 (0.54-1.52) | 0.50 (0.21-1.21) | 0.71 |

Abbreviations: HR (hazard ratio), CI (confidence interval)

[a] Overall social distancing grades are denoted as Poor (F grade), Fair (D grade), Good (C grade), and Excellent (A+B grade). Overall social grade categories (A, B, C, D, and F) are provided by Unacast.

[b] P value for trend is calculated using the median value of each category as a continuous variable.

[c] Model 1 was stratified by age (<25, 25-34, 35-44, 45-54, 55-64, or ≥65), state, and calendar date at study entry.

[d] Model 2 was stratified by age (<25, 25-34, 35-44, 45-54, 55-64, ≥65), state, and calendar date at study entry and further adjusted for race (white, black, Asian, or other), sex (male or female), population density of residence (quartiles), current smoking, frontline healthcare worker, interaction with suspected or documented Covid-19, history of diabetes, heart disease, lung disease, and kidney disease (each yes or no).

**Supplemental Table 4. Risk of testing positive for COVID-19 according to personal use of a face mask**

| | | Personal use of a face mask [a] | |
|---|---|---|---|
| | | Never | Ever [b] |
| Overall social distancing | | | |
| | No. of Case/ Person-time (days) | 57/2660436 | 7/1325035 |
| | Model 1 HR (95% CI) [c] | 1 [Reference] | 0.25 (0.11-0.58) |
| | Model 2 HR (95% CI) [d] | 1 [Reference] | 0.27 (0.12-0.61) |
| Poor (F) [e] | | | |
| | No. of Case/ Person-time (days) | 48/1980588 | 6/1002943 |
| | Model 1 HR (95% CI) | 1 [Reference] | 0.24 (0.10-0.59) |
| | Model 2 HR (95% CI) | 1 [Reference] | 0.25 (0.10-0.62) |

Abbreviations: HR (hazard ratio), CI (confidence interval)

[a] Use of a face mask was collected from 139,690 participants beginning on June 12, 2020 based on the query “In the last week, did you wear a face mask when outside the house?”.

[b] Ever wearing a face mask includes sometimes, most of the time, or always wearing a face mask as a time-varying variable.

[c] Model 1 was stratified by age (<25, 25-34, 35-44, 45-54, 55-64, or ≥65), state, and calendar date at study entry.

[d] Model 2 was stratified by age (<25, 25-34, 35-44, 45-54, 55-64, ≥65), state, and calendar date at study entry and further adjusted for race (white, black, Asian, or other), sex (male or female), population density (quartiles), current smoking, frontline healthcare worker, interaction with suspected or documented Covid-19, history of diabetes, heart disease, lung disease, and kidney disease (each yes or no).

[e] Only poor(F) social distance grade group is demonstrated due to a limited number of participants in Fair (D grade), Good (C grade), and Excellent (A+B grade).

**Supplemental Table 5. Risk of predicted COVID-19 according to overall social distancing grade within Rt subgroups**

| | Overall social distancing grade [a] | | | | |
|---|---|---|---|---|---|
| Effective reproductive number (Rt) | Poor (F) | Fair (D) | Good (C) | Excellent (A/B) | P value for Trend [b] |
| ≤1.0 | | | | | |
| No. of Case/ Person-time (days) | 482/1807347 | 729/2282849 | 765/1940771 | 82/248248 | |
| Model 1 HR (95% CI) [c] | 1 [Reference] | 0.86 (0.74-1.00) | 0.77 (0.64-0.92) | 0.62 (0.46-0.84) | <.001 |
| Model 2 HR (95% CI) [d] | 1 [Reference] | 0.88 (0.75-1.02) | 0.79 (0.66-0.96) | 0.64 (0.48-0.87) | 0.004 |
| >1.0 | | | | | |
| No. of Case/ Person-time (days) | 1056/2842727 | 728/1397419 | 587/793043 | 59/91364 | |
| Model 1 HR (95% CI) | 1 [Reference] | 0.84 (0.72-0.97) | 0.82 (0.68-0.99) | 0.76 (0.54-1.05) | 0.04 |
| Model 2 HR (95% CI) | 1 [Reference] | 0.85 (0.73-0.98) | 0.83 (0.69-1.00) | 0.82 (0.59-1.14) | 0.08 |

Abbreviations: HR (hazard ratio), CI (confidence interval)

[a] Overall social distancing grades are denoted as Poor (F grade), Fair (D grade), Good (C grade), and Excellent (A+B grade). Overall social grade categories (A, B, C, D, and F) are provided by Unacast.

[b] P value for trend is calculated using the median value of each category as a continuous variable.

[c] Model 1 was stratified by age (<25, 25-34, 35-44, 45-54, 55-64, or ≥65), state, and calendar date at study entry.

[d] Model 2 was stratified by age (<25, 25-34, 35-44, 45-54, 55-64, ≥65), state, and calendar date at study entry and further adjusted for race (white, black, Asian, or other), sex (male or female), population density of residence (quartiles), current smoking, frontline healthcare worker, interaction with suspected or documented Covid-19, history of diabetes, heart disease, lung disease, and kidney disease (each yes or no).

**Supplemental Table 6. Risk of predicted COVID-19 according to Personal use of a face mask within Rt subgroups**

| | | Personal use of a face mask [a] | |
|---|---|---|---|
| Effective reproductive number (Rt) | | **Never** | **Ever** [b] |
| ≤1.0 | | | |
| | No. of Case/ Person-time (days) | 195/705433 | 53/333864 |
| | Model 1 HR (95% CI) [c] | 1 [Reference] | 0.33 (0.22-0.49) |
| | Model 2 HR (95% CI) [d] | 1 [Reference] | 0.33 (0.22-0.50) |
| >1.0 | | | |
| | No. of Case/ Person-time (days) | 623/1789196 | 215/856702 |
| | Model 1 HR (95% CI) | 1 [Reference] | 0.34 (0.28-0.42) |
| | Model 2 HR (95% CI) | 1 [Reference] | 0.35 (0.28-0.43) |

Abbreviations: HR (hazard ratio), CI (confidence interval)

[a] Use of a face mask was collected from 139,690 participants beginning on June 12, 2020 based on the query "In the last week, did you wear a face mask when outside the house?".

[b] Ever wearing a face mask includes sometimes, most of the time, or always wearing a face mask as a time-varying variable.

[c] Model 1 was stratified by age (<25, 25-34, 35-44, 45-54, 55-64, or ≥65), state, and calendar date at study entry.

[d] Model 2 was stratified by age (<25, 25-34, 35-44, 45-54, 55-64, ≥65), state, and calendar date at study entry and further adjusted for race (white, black, Asian, or other), sex (male or female), population density (quartiles), current smoking, frontline healthcare worker, interaction with suspected or documented Covid-19, history of diabetes, heart disease, lung disease, and kidney disease (each yes or no).
